# Trends in the burden of sickle cell disease in Sierra Leone, 1990–2023: a Global Burden of Disease analysis

**DOI:** 10.64898/2025.12.01.25341432

**Authors:** Monalisa M.J. Faulkner, Fatima Jalloh, Foray Mohamed Foray, Sahr L. Gborie, Mohamed B. Jalloh

## Abstract

Sickle cell disease is common in Sierra Leone, yet its long-term population burden is uncertain. We analyzed Global Burden of Disease 2023 estimates for sickle cell disorders in Sierra Leone from 1990 through 2023. We obtained prevalence, deaths, and disability adjusted life years (DALYs), calculated age standardized rates per 100 000, modeled temporal trends with log linear regression, decomposed mortality changes into effects of population growth and rate change, and assessed age specific and sex specific patterns in 2023. Sickle cell prevalent cases increased from 48 689 (95% UI, 42 588 to 56 140) in 1990 to 90 498 (95% UI, 78 126 to 105 815) in 2023, and deaths rose from 408 (95% UI, 288 to 579) to 635 (95% UI, 438 to 862). DALYs increased from 32 518 (95% UI, 23 446 to 45 336) to 50 608 (95% UI, 36 331 to 66 317). Over the same period, age standardized mortality declined from 10.2 to 7.9 per 100 000 (APC, −0.46%; 95% CI, −0.64 to −0.29) and age standardized DALYs from 811.5 to 631.5 per 100 000 (APC, −0.43%; 95% CI, −0.60 to −0.25). In 2023, children younger than 5 years accounted for 153 deaths (95% UI, 93 to 230; 24.1% of all deaths), and persons younger than 20 years for 314 deaths (95% UI, 203 to 446; 49.5%). Males and females had similar prevalence (45 442 vs. 45 056 cases), but males had 371 deaths and 28 855 DALYs versus 264 deaths and 21 753 DALYs in females. Decomposition indicated that population growth alone would have increased deaths by 179.5%, whereas rate reductions offset this by 79.5%. The burden of sickle cell disease in Sierra Leone has grown in absolute terms, driven mainly by rapid population expansion, while age standardized mortality and DALY rates have declined. Concentrated losses in children and young adults and persistent male excess mortality highlight the need to scale up newborn screening and early comprehensive care, expand access to hydroxyurea and infection prevention, and strengthen resilient sickle cell services.

## Introduction

Sickle cell disease is among the most common severe inherited blood disorders worldwide and is a major cause of premature death and disability in low- and middle-income countries (LMICs)(1,2). Global modeling indicates that hundreds of thousands of infants are born each year with sickle cell disease and that this number is likely to rise as populations grow and child survival improves(3,4). The burden is concentrated in sub-Saharan Africa (SSA), which accounts for more than three quarters of prevalent cases; in many settings in this region, between one half and nine tenths of affected children die before 5 years of age in the absence of systematic newborn screening and comprehensive care(3–5). Although effective interventions, including early diagnosis, infection prophylaxis, hydroxyurea therapy, and structured chronic care, are available, access to these services remains limited in many high burden countries(2,5)

International agencies have identified sickle cell disease as a public health priority. In 2006 the World Health Assembly urged member states to strengthen prevention and management of sickle cell disease, and subsequent regional frameworks have called for integration of sickle cell services into primary care and maternal and child health programs(6,7). Despite these commitments, reports from across Africa indicate that most countries still lack population-based surveillance, newborn screening programs, and robust data on long term outcomes(5,8)

Sierra Leone is thought to have one of the highest burdens of sickle cell disease in West Africa. Trait frequencies have been estimated at 20 to 25%, and modeling studies suggest several thousand affected births each year(9–11). Hospital based reports show that sickle cell disease contributes substantially to anemia, hospital admission, and child mortality, yet specialized services are scarce and newborn screening has only recently been piloted(9,12,13). Recent expert commentary has characterized sickle cell disease as a neglected public health problem in Sierra Leone and has called for quantitative estimates of its population level burden to guide policy and program development(9,13,14)

The Global Burden of Disease (GBD) collaboration provides standardized, internally consistent estimates of mortality, years of life lost (YLL), years lived with disability (YLD), and disability adjusted life years (DALYs) for hundreds of conditions across countries and over time(15,16). Recent GBD analyses have quantified the global and regional burden of sickle cell disease, but detailed country specific profiles for high burden settings remain limited(1). In particular, no previous study has systematically described long term trends in sickle cell disease burden, age and sex patterns, and drivers of change in Sierra Leone using contemporary GBD data.

In this study, we used GBD 2023 estimates to describe temporal trends from 1990 through 2023 in prevalence, mortality, years of life lost, years lived with disability, and disability adjusted life years attributed to sickle cell disease in Sierra Leone. We further characterized the distribution of burden by age group and sex and decomposed changes in deaths into components attributable to population growth and to changes in mortality rates. Our objective was to generate decision grade evidence on the evolving burden of sickle cell disease in Sierra Leone to inform national policy, service planning, and future research.

## Methods

### Study design and data source

We conducted a secondary analysis of estimates from the GBD 2023 study, produced by the Institute for Health Metrics and Evaluation (IHME). The GBD enterprise applies a standardised cause hierarchy, systematic data synthesis and Bayesian meta-regression (DisMod-MR 2.1) to produce internally consistent estimates of incidence, prevalence, mortality, YLLs, YLDs and DALYs across 204 countries and territories(16). For this analysis, we extracted all available data for Sierra Leone for sickle cell disorders (GBD cause ID 615) for calendar years 1990 through 2023. No modification of the underlying GBD models was undertaken; our work represents a country focused re analysis and interpretation of the official GBD 2023 results for sickle cell disorders. The overall GBD methods have been reported in detail elsewhere and include extensive procedures to correct bias in primary data, reassign nonspecific “garbage codes” in vital registration, and carry forward parameter uncertainty through posterior simulation(17)

### Measures and metrics

We examined five standard GBD burden measures. Prevalence was defined as the number of individuals living with sickle cell disease in a given year. Deaths captured the number of deaths attributable to sickle cell disorders as the underlying cause. YLDs quantified time spent in less than full health due to non-fatal complications of sickle cell disease, weighted by disability severity. YLLs reflected premature mortality and were calculated as the product of cause specific deaths and the GBD reference life expectancy at age of death. DALYs were derived as the sum of YLDs and YLLs and represent the total number of years of healthy life lost due to sickle cell disease.

For each measure, we analysed both absolute counts and age standardized rates per 100 000 population. Age standardized rates were computed by GBD using the standard GBD global age structure, enabling comparison across time and across populations with differing age distributions. All reported estimates are accompanied by 95% uncertainty intervals (UIs) derived from 1 000 posterior draw level simulations, which propagate uncertainty from input data, data adjustments, and model parameters through to the final estimates.

### Stratification

We summarised burden for each calendar year from 1990 to 2023. To describe the age pattern of sickle cell disease in a way that is relevant to policy, we used three overlapping age strata: younger than 5 years, younger than 20 years, and 15 to 49 years, in addition to all ages combined. These categories were treated as cumulative and overlapping and therefore were not summed. All metrics were stratified by sex (male, female, and both sexes combined).

### Statistical analysis

To characterise temporal trends, we modelled annual age standardized rates using log linear regression of the form

ln(rate) = β₀ + β₁ × (calendar year).

The annual percent change (APC) in the age standardized rate was calculated as APC = [exp(β₁) − 1] × 100.

95% confidence intervals (CIs) for the APC were obtained from the standard error of β₁, assuming approximate normality of the estimator. A two-sided alpha level of 0.05 was used to define statistical significance.

We decomposed the change in the number of deaths between 1990 and 2023 into two components: the contribution of population growth and the contribution of changes in age standardized mortality rates. For each year, we inferred the effective population size from the relationship

population = (deaths / mortality rate) × 100 000.

We then estimated the number of deaths that would have occurred in 2023 if age standardized mortality rates had remained at their 1990 values, but the population had grown to its 2023 size. The difference between this counterfactual number and the observed number of deaths in 1990 was attributed to population growth, whereas the remaining difference between this counterfactual and the observed deaths in 2023 was attributed to changes in mortality rates.

To explore sex-based inequities, we calculated male to female ratios for each burden metric in 2023 within each age stratum. Ratios greater than 1.20 or less than 0.83 were prespecified as indicating substantial disparity that would merit programmatic or policy attention, recognising that smaller deviations from unity may still be important in clinical settings.

### Software and data visualisation

All analyses were conducted in Python 3.12 using pandas (data manipulation), NumPy (numerical operations), SciPy (statistical tests), matplotlib and seaborn (visualization). Graphical displays present point estimates with 95% UIs shown as shaded bands for time series plots and as error bars for bar charts.

### Ethical considerations

This study used only publicly available, de identified, aggregate GBD 2023 estimates. No individual level data were accessed, and no linkages to identifiable information were performed. In accordance with prevailing guidance for analyses based solely on public domain aggregate data, institutional review board approval was not required.

## Results

### Overall burden and temporal trends, 1990 to 2023

Between 1990 and 2023, the absolute burden of sickle cell disease in Sierra Leone increased substantially across all measures. The number of prevalent cases rose from 48,689 (95% UI, 42,588 to 56,140) in 1990 to 90,498 (95% UI, 78,126 to 105,815) in 2023, representing an 85.9% increase. Annual deaths attributable to sickle cell disease increased from 408 (95% UI, 288 to 579) to 635 (95% UI, 438 to 862), a 55.7% increase. DALYs rose from 32,518 (95% UI, 23,446 to 45,336) to 50,608 (95% UI, 36,331 to 66,317), reflecting a 55.6% increase (**Table 1**, **Figure 1**).

**Figure 1.**
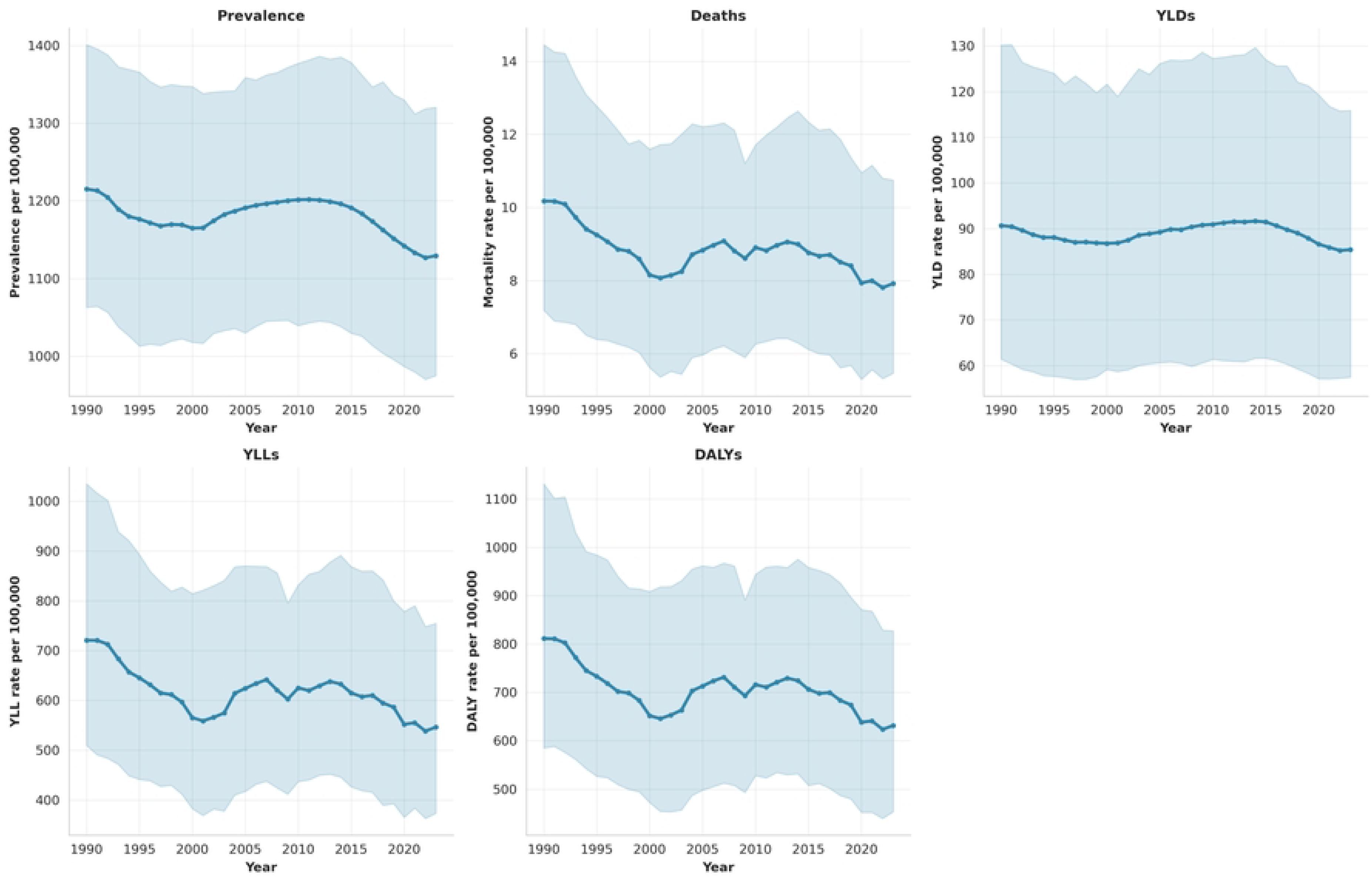
Temporal trends in sickle cell disease burden in Sierra Leone, 1990–2023. Absolute counts and age-standardized rates per 100,000 population for prevalence, deaths, years lived with disability (YLDs), years of life lost (YLLs), and disability-adjusted life-years (DALYs).

**Table 1:** Burden of sickle cell disease in Sierra Leone, 1990 and 2023, and percentage change in counts and age-standardized rates.

| Measure | 1990 Count<br>(95% UI) | 2023<br>Count<br>(95%<br>UI) | Percentage<br>change in<br>counts | 1990 Age-standardized rate<br>per 100,000 (95%<br>UI) | 2023 Age-standardized rate<br>per 100,000 (95%<br>UI) | Percentage change<br>in age-standardized rates |
| --- | --- | --- | --- | --- | --- | --- |
| Prevalence | 48,689<br>(42,588–<br>56,140) | 90,498<br>(78,126–<br>105,815) | 85.9% | 1215.0 (1062.7–<br>1400.9) | 1129.3 (974.9–<br>1320.4) | –7.1% |
| Deaths | 408 (288–<br>579) | 635<br>(438–<br>862) | 55.7% | 10.2 (7.2–14.4) | 7.9 (5.5–10.8) | –22.1% |
| YLDs | 3,634<br>(2,462–<br>5,217) | 6,843<br>(4,605–<br>9,286) | 88·3% | 90·7 (61·4–<br>130·2) | 85·4 (57·5–<br>115·9) | –5·8% |
| YLLs | 28,884<br>(20,422–<br>41,472) | 43,765<br>(29,905–<br>60,483) | 51·5% | 720·8 (509·6–<br>1034·9) | 546·1 (373·2–<br>754·7) | –24·2% |
| DALYs | 32,518<br>(23,446–<br>45,336) | 50,608<br>(36,331–<br>66,317) | 55·6% | 811·5 (585·1–<br>1131·3) | 631·5 (453·4–<br>827·5) | –22·2% |

Data are counts or age-standardized rates per 100,000 population. UI=uncertainty interval.
YLDs=years lived with disability. YLLs=years of life lost. DALYs=disability-adjusted life-years.

Despite these increases in absolute burden, age-standardized rates declined significantly over the study period. The age-standardized prevalence rate decreased from 1215.0 per 100,000 population in 1990 to 1129.3 per 100,000 in 2023 (−7.1%). More notably, the age-standardized mortality rate declined from 10.2 per 100,000 to 7.9 per 100,000 (−22.1%), and the age-standardized DALY rate fell from 811.5 per 100,000 to 631.5 per 100,000 (−22.2%). YLLs declined by 24.2%, from 720.8 to 546.1 per 100,000 (**Table 1**, **Figure 1**).

Log-linear regression analysis revealed statistically significant annual declines in age-standardized rates for mortality (annual percent change [APC], −0.46%; 95% CI, −0.64 to −0.29; P<0.0001), DALYs (APC, −0.43%; 95% CI, −0.60 to −0.25; P=0.0001), and YLLs (APC, −0.48%; 95% CI, −0.68 to −0.29; P<0.0001). The decline in prevalence rate was modest (APC, −0.10%; 95% CI, −0.16 to −0.04; P=0.003), while the rate of years lived with disability (YLDs) showed no significant trend (APC, −0.01%; 95% CI, −0.08 to 0.07; P=0.82) (**Table 2)**.

**Table 2:** Annual percentage change in age-standardized rates of sickle cell disease burden, 1990–2023.

| Measure | Annual percentage change (95% CI) | p value | R<sup>2</sup> |
| --- | --- | --- | --- |
| Prevalence | −0.10 (−0.16 to −0.04) | 0.003 | 0.243 |
| Deaths | −0.46 (−0.64 to −0.29) | <0.0001 | 0.454 |
| YLDs | −0.01 (−0.08 to 0.07) | 0.823 | 0.002 |
| YLLs | −0.48 (−0.68 to −0.29) | <0.0001 | 0.419 |
| DALYs | −0.43 (−0.60 to −0.25) | <0.0001 | 0.401 |

Annual percentage change calculated from age-standardised rates. CI=confidence interval.
YLDs=years lived with disability. YLLs=years of life lost. DALYs=disability-adjusted life-years.

### Age distribution of disease burden in 2023

In 2023, the burden of sickle cell disease was concentrated among children and young adults. Among children younger than 5 years, there were 153 deaths (95% UI, 93 to 230), accounting for 24.1% of all sickle cell disease deaths. Deaths among persons younger than 20 years totaled 314 (95% UI, 203 to 446), representing 49.5% of all deaths. The highest absolute death burden occurred in the 15-to-49-year age group (380 deaths; 95% UI, 264 to 512), which comprised 59.9% of total mortality (**Table 3**, **Figure 2**).

**Figure 2.**
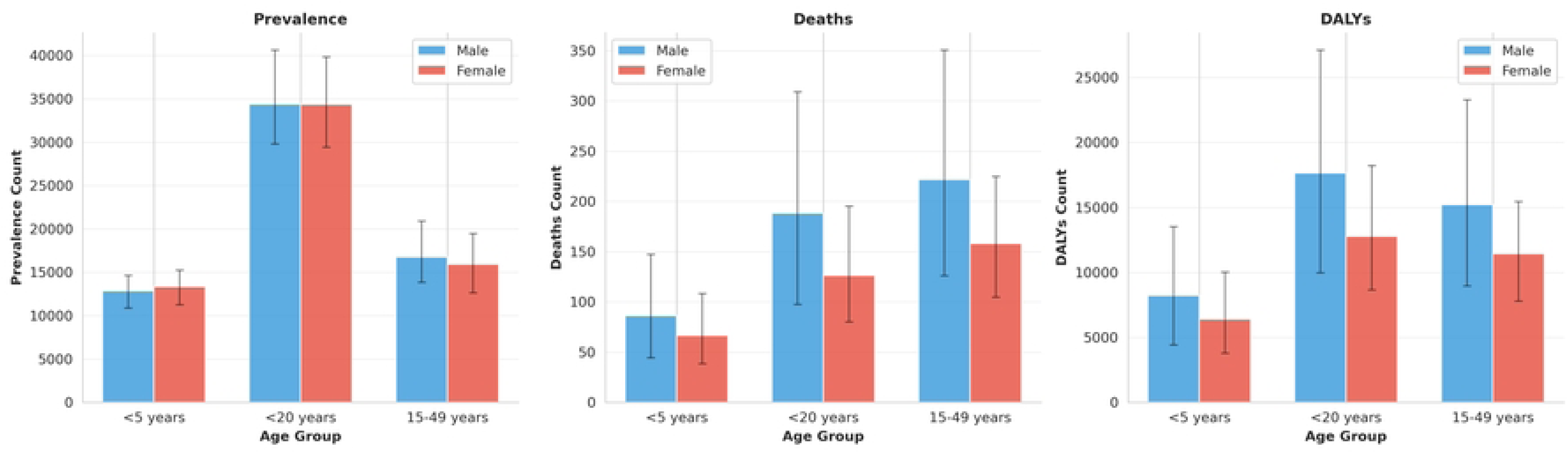
Age and sex distribution of sickle cell disease burden in 2023. Prevalence, deaths, and disability-adjusted life-years (DALYs) due to sickle cell disease in Sierra Leone by age group and sex.

**Table 3:** Age and sex distribution of sickle cell disease burden in 2023.

| Measure | Age group | Both sexes (95% UI) | Male (95% UI) | Female (95% UI) | Male-to-female ratio |
| --- | --- | --- | --- | --- | --- |
| Prevalence |  |  |  |  |  |
|  | <5 years | 26,189 (22,349–29,463) | 12,863 (10,877–14,630) | 13,326 (11,245–15,235) | 0.97 |
|  | <20 years | 68,769 (59,528–79,803) | 34,399 (29,789–40,604) | 34,369 (29,415–39,833) | 1.00 |
|  | 15–49 years | 32,711 (26,542–39,486) | 16,779 (13,820–20,907) | 15,932 (12,628–19,464) | 1.05 |
| Deaths |  |  |  |  |  |
|  | <5 years | 153 (93–230) | 86 (44–147) | 67 (38–108) | 1.30 |
|  | <20 years | 314 (203–446) | 188 (97–309) | 126 (80–195) | 1.49 |
|  | 15–49 years | 380 (264–512) | 222 (126–350) | 158 (105–225) | 1·40 |
| DALYs |  |  |  |  |  |
|  | <5 years | 14,623 (9,127–21,750) | 8,220 (4,417–13,506) | 6,402 (3,786–10,014) | 1·28 |
|  | <20 years | 30,437 (21,303–40,768) | 17,657 (9,978–27,098) | 12,780 (8,638–18,217) | 1·38 |
|  | 15–49 years | 26,647 (19,205–34,951) | 15,219 (8,963–23,271) | 11,427 (7,792–15,444) | 1·33 |

Data are counts. UI=uncertainty interval. DALYs=disability-adjusted life-years.

Prevalence was highest among persons younger than 20 years (68,769 cases; 95% UI, 59,528 to 79,803), with children younger than 5 years accounting for 26,189 cases (95% UI, 22,349 to 29,463). The reproductive-age population (15 to 49 years) included 32,711 persons living with sickle cell disease (95% UI, 26,542 to 39,486). DALYs followed a similar age pattern, with 30,437 DALYs (95% UI, 21,303 to 40,768) among persons younger than 20 years and 26,647 DALYs (95% UI, 19,205 to 34,951) among those 15 to 49 years of age (**Table 3**).

### Sex differences in disease burden

Males experienced substantially higher mortality and morbidity than females despite nearly equal disease prevalence. In 2023, the male-to-female prevalence ratio was 1.01 (45,442 vs. 45,056 cases), indicating equal disease distribution by sex. However, males had 40% more deaths than females (371 vs. 264; male-to-female ratio, 1.40) and 33% more DALYs (28,855 vs. 21,753; male-to-female ratio, 1.33). YLLs were similarly disproportionate (25,535 vs. 18,230; male-to-female ratio, 1.40), whereas YLDs showed a slight female predominance (3,320 vs. 3,523; male-to-female ratio, 0.94) (**Figure 2**).

### Decomposition of mortality trends

The observed increase of 227 deaths from 1990 to 2023 was decomposed into two principal components: population growth and rate change (**Figure 3**). Sierra Leone’s population doubled during the study period, from approximately 4.0 million to 8.0 million. If the 1990 age-standardized mortality rate had remained constant, this population growth alone would have produced 408 additional deaths (179.5% of the observed change). However, improvements in sickle cell disease care and survival reduced the age-standardized mortality rate by 22.1%, preventing an estimated 181 deaths (−79.5% of the observed change). Thus, without population growth, sickle cell disease mortality would have declined by 44%. The competing effects of population expansion and declining mortality rates resulted in the net increase of 227 deaths.

**Figure 3.**
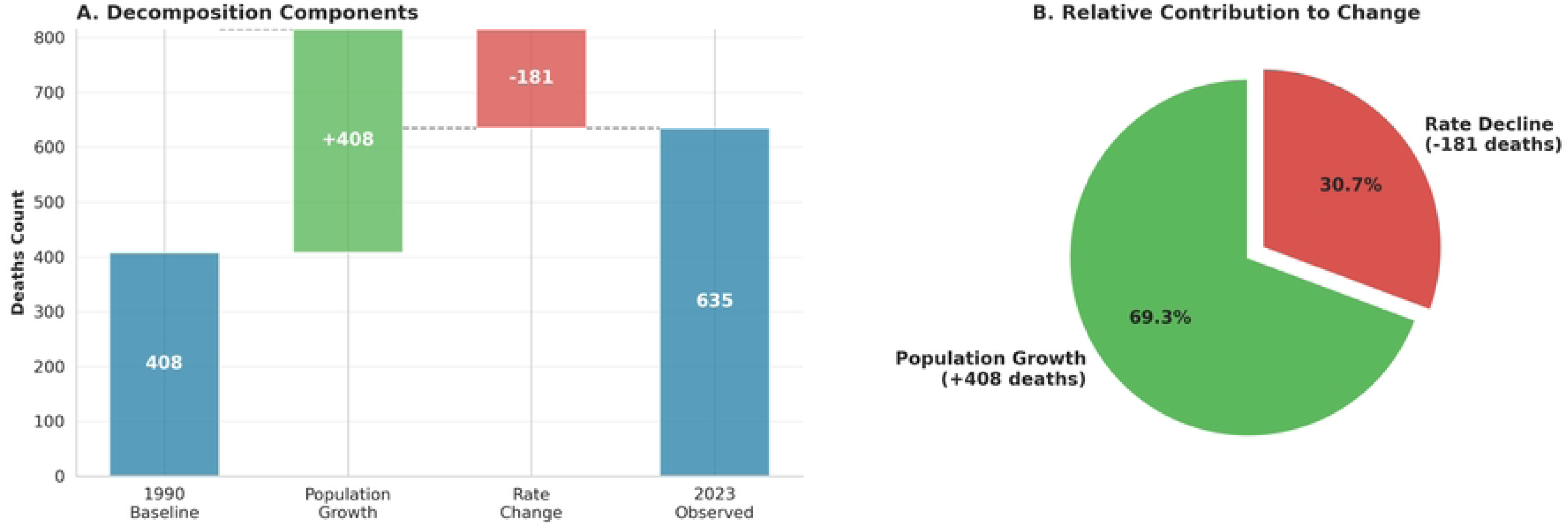
Decomposition of the change in sickle cell disease deaths between 1990 and 2023. Contributions of population growth, population ageing, and changes in age-specific mortality rates to the net change in the number of deaths.

Age-stratified analyses revealed persistent male excess mortality across all age groups. Among children younger than 5 years, the male-to-female mortality ratio was 1.30; among persons younger than 20 years, the ratio was 1.49; and among those 15 to 49 years of age, the ratio was 1.40. This consistent pattern suggests potential sex-based differences in disease severity, health care access, or health-seeking behavior (**Figure 2**, **Figure 4**).

**Figure 4.**
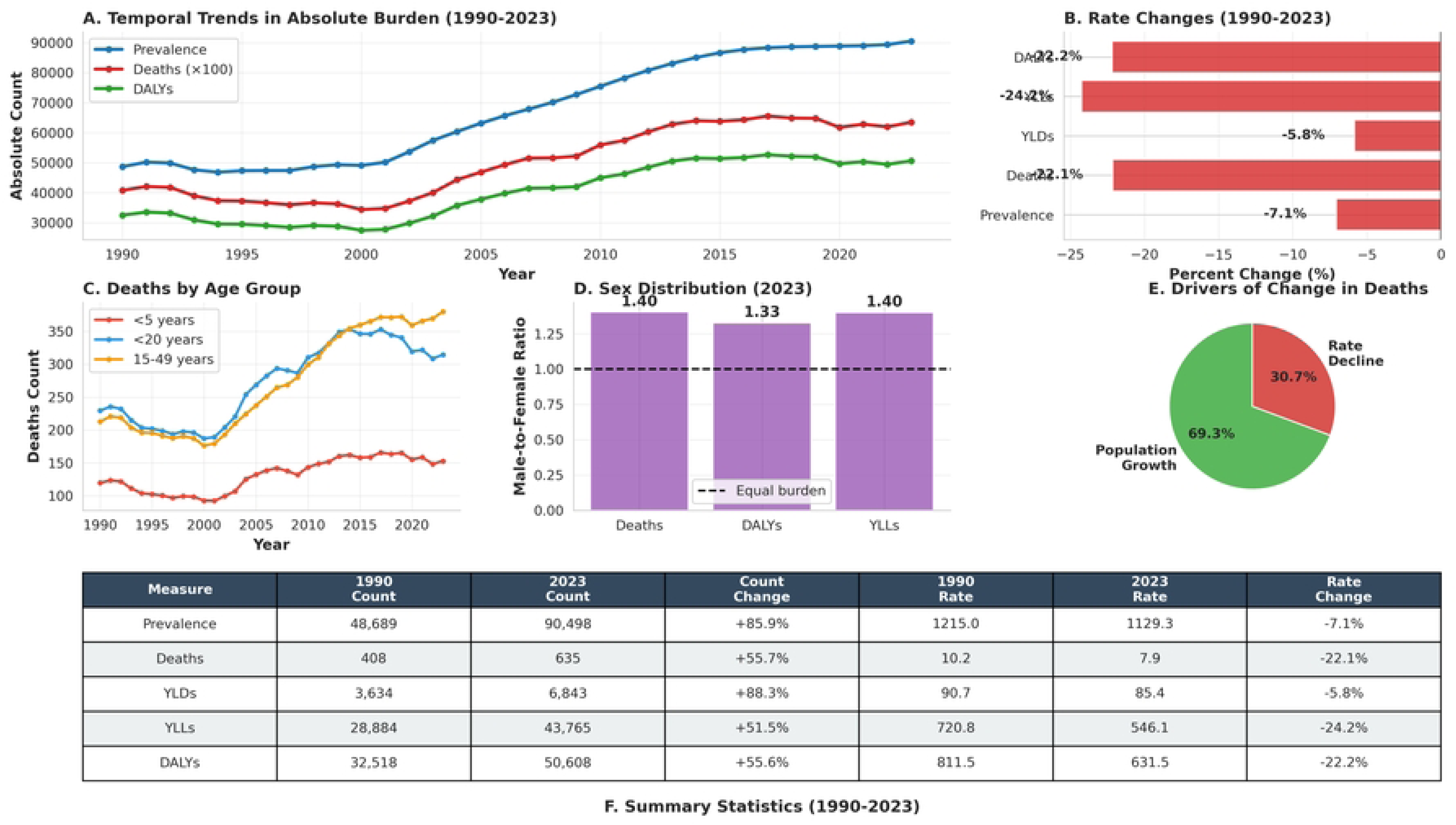
Comprehensive summary of sickle cell disease burden in Sierra Leone, 1990-2023. Relative contributions of major age groups and sexes to total prevalence, deaths, and disability-adjusted life-years (DALYs) from 1990 to 2023.

## Discussion

In this nationwide, three-decade analysis of GBD data for Sierra Leone, we found that the absolute burden of sickle cell disease increased substantially between 1990 and 2023, largely because of population growth, whereas age standardized mortality and DALY rates declined. In decomposition analyses, deaths attributable to sickle cell disease would have decreased in the absence of population expansion, consistent with gradual gains in survival. The burden remained concentrated in children and young people, with nearly half of deaths occurring before 20 years of age, and males had disproportionately higher mortality and years of life lost despite similar prevalence (**Figure 4**). These patterns are concordant with the broader global assessment of sickle cell disease in the GBD 2021 analysis, which showed high mortality before 5 years of age in SSA together with declining rates in several high incidence countries (1).

The combination of rising case numbers and falling age standardized rates reflects a characteristic epidemiologic transition in sickle cell disease. Regional and multi-country analyses have documented substantial excess mortality in early childhood in SSA, with under 5 mortality among children with sickle cell anemia several times higher than in unaffected peers (5,11). In parallel, the GBD sickle cell disease study showed that, although cause specific mortality is highest in young children, rates have begun to fall where core components of comprehensive care are available (1,18). Our findings in Sierra Leone align with this pattern: population growth and improved survival among affected children together yield more people living with sickle cell disease, even as age standardized mortality and DALY rates decline.

The age distribution of burden in our analysis, with high prevalence in children, significant mortality before 5 years of age, and large DALY losses among adolescents and young adults, is consistent with birth cohort and registry data. In the Jamaican cohort, followed from birth into adulthood, simple protocolized interventions after newborn diagnosis, including penicillin prophylaxis, vaccination, and parent education on splenic sequestration, substantially reduced early mortality (19). The Lancet Haematology Commission on sickle cell disease argued that extending such packages at scale in high incidence countries is central to shifting the burden from early childhood death toward chronic, manageable disease (18). Emerging African implementation platforms, including the Consortium on Newborn Screening in Africa (CONSA), are now providing prospective evidence that integration of newborn screening with standardized early care can improve survival in high incidence settings (20). Our observation that absolute deaths increased primarily because more children with sickle cell disease survived long enough to remain at risk is consistent with the dynamics reported in these programmatic cohorts.

The marked excess in mortality, years of life lost, and DALYs among males in Sierra Leone, despite similar prevalence in males and females, is in keeping with reports of sex-based differences in clinical severity and outcomes. In the Sickle Cell Disease Implementation Consortium, males had higher odds of several life-threatening complications and chronic end organ damage, even after adjustment for age and genotype (21). Other observational studies have suggested that lower fetal hemoglobin levels, more intense hemolysis, and differences in nitric oxide bioavailability may contribute to poorer outcomes in males, while gendered patterns in health seeking behavior may amplify biological risk (18,22,23). In Sierra Leone, these findings support sex responsive risk stratification, with particular attention to adolescent and young adult males for retention in care, screening for complications, and adherence support.

Several system level and intervention related factors may have contributed to the favorable rate trends. Vaccination and infection prevention are central. Pneumococcal conjugate vaccines have reduced invasive pneumococcal disease in children, including those with sickle cell disease, although serotype replacement and residual risk persist and case fatality among affected children remains high (24,25). These observations reinforce the importance of strict penicillin prophylaxis and completion of expanded pneumococcal vaccination schedules, including polysaccharide boosters where available, for all children with sickle cell disease. Disease modifying therapy has also become increasingly feasible in African settings. In a multi-country trial in SSA, hydroxyurea at moderate doses was safe and significantly reduced vaso-occlusive pain, acute chest syndrome, transfusion requirements, and hospitalization in children with sickle cell anemia (26,27). Long term data from the Realizing Effectiveness across Continents with Hydroxyurea (REACH) trial indicate that dose optimization toward the maximum tolerated dose is feasible and associated with sustained benefits, including fewer severe events and improved growth (28,29). Scaling access to hydroxyurea within simplified monitoring algorithms is therefore a realistic strategy to further reduce sickle cell disease mortality and DALYs in Sierra Leone.

Health system shocks over the study period may have attenuated these gains. The 2014 to 2015 Ebola epidemic in Sierra Leone led to sharp declines in the use of maternal and child health services and was estimated to increase under 5 mortality nationally by approximately one fifth (30,31). Children with chronic conditions such as sickle cell disease are particularly vulnerable to interruptions in prophylaxis, transfusion support, and acute care during such crises. Although our time series analysis cannot isolate the transient effects of Ebola on sickle cell disease outcomes, the epidemic underscores the importance of resilient, decentralized systems for follow up and drug supply that can maintain continuity of care during future shocks.

The policy implications of these findings are direct. First, newborn screening for sickle cell disease should be integrated into routine immunization and child health platforms, using point of care diagnostics where laboratory capacity is limited, as shown in Nigeria and within CONSA (5,20). Early identification needs to be paired with a standardized, low cost package of care that includes penicillin prophylaxis, folate supplementation, malaria prevention, and on schedule Haemophilus influenzae type b and pneumococcal vaccination. Second, national protocols should ensure completion of pneumococcal vaccination with attention to nonvaccine serotypes, reliable penicillin supply, and rapid evaluation of febrile illness in children with sickle cell disease (24,25). Third, access to hydroxyurea for children and adolescents with sickle cell anemia should be expanded, with dose titration guided by simple laboratory markers, building on African trial experience (26). Finally, given the persistent male disadvantage, adolescent and young adult males should be prioritized for retention in care, screening for complications, and tailored adherence and mental health support (32). Building robust supply chains and community based follow up will be essential to sustain these services during future outbreaks or other system disruptions.

### Strengths and limitations

This study has several strengths. We used standardized GBD 2023 estimates over more than three decades, which allowed consistent comparison of counts and rates, and we explicitly decomposed mortality trends into components attributable to population growth and to changes in mortality rates. This approach clarified that population expansion masked real gains in reducing mortality risk. Age and sex disaggregation revealed actionable inequities, including concentrated losses in young children and persistent excess mortality among males, findings that align with multi-country analyses and Commission level syntheses (1,18).

Important limitations arise from reliance on modeled estimates rather than nationwide registries. For Sierra Leone, GBD estimates depend on sparse primary data, and under ascertainment of early childhood deaths is a recognized challenge in sickle cell disease epidemiology. Our analysis cannot attribute observed declines in rates to specific interventions or programs, and the effects of major health system shocks, including Ebola, are difficult to separate from longer term trends. These constraints highlight the need for a national sickle cell disease registry in Sierra Leone, ideally linked to newborn screening and longitudinal follow up, to directly measure survival, treatment coverage, and cause specific mortality.

## Conclusion

From 1990 through 2023, the absolute burden of sickle cell disease in Sierra Leone increased, whereas age standardized mortality and DALY rates declined, indicating incremental improvements in survival that did not fully offset rapid population growth. The concentration of deaths in children and adolescents and the consistent excess mortality and years of life lost among males define an urgent agenda: expand newborn screening with early comprehensive care, ensure robust infection prevention and pneumococcal vaccination with penicillin adherence, broaden access to hydroxyurea with dose optimization, and strengthen resilient child health and hematology services that can withstand future health system shocks.

## Abbreviations

APC: Annual percent change
CI: Confidence interval
CONSA: Consortium on Newborn Screening in Africa
DALY: Disability-adjusted life year
GBD: Global Burden of Disease
ICD: International Classification of Diseases
IHME: Institute for Health Metrics and Evaluation
LMIC: Low- and middle-income country
NCD: Non-communicable disease
REACH: Realizing Effectiveness across Continents with Hydroxyurea
SSA: Sub-Saharan Africa
WHO: World Health Organization
YLD: Years lived with disability
YLL: Years of life lost

## Declarations

### Consent for publication

Not applicable

### Data availability

GBD 2023 summary estimates are publicly available through the Institute for Health Metrics and Evaluation data portal. Available from https://vizhub.healthdata.org/gbd-results/.

### Competing interests

The authors declare no competing interests.

### Funding

This work did not receive any funding.

### Authors’ contributions

MBJ conceived and designed the study, curated the data, and led the analysis. MBJ and MMJF developed the analytic approach. MMJF, FJ, FMF, and SLG contributed to the interpretation of the findings and to the contextualisation of the results within Sierra Leone’s health system. MBJ and MMJF drafted the first version of the manuscript. All authors critically reviewed the manuscript for important intellectual content and approved the final version for submission.

## Acknowledgments

We acknowledge the Institute for Health Metrics and Evaluation and GBD collaborators for producing and sharing the 2023 estimates.

## Notes

### Competing Interest Statement

The authors have declared no competing interest.

